# Estimated changes in free sugar consumption one year after the UK Soft drinks industry levy came into force: controlled interrupted time series analysis of the National Diet and Nutrition Survey (2011-2019)

**DOI:** 10.1101/2023.06.26.23291902

**Authors:** Nina T Rogers, Steven Cummins, Catrin P Jones, Oliver Mytton, Mike Rayner, Harry Rutter, Martin White, Jean Adams

## Abstract

**Background:** The UK soft drinks industry levy (SDIL) was announced in March 2016 and implemented in April 2018, encouraging manufacturers to reduce the sugar content of soft drinks. This is the first study to investigate changes in individual-level consumption of free sugars in relation to the SDIL.

**Methods:** We used controlled interrupted time series (2011-2019) to explore changes in consumption of free sugars in the whole diet and from soft drinks alone, 11 months post-SDIL implementation in a nationally representative sample of adults (>18y;n=7,999) and children (1.5-19y;n=7656) drawn from the UK National Diet and Nutrition Survey. Estimates were based on differences between observed data and a counterfactual scenario of no SDIL announcement/implementation. Models included protein consumption (control) and accounted for autocorrelation.

**Results:** Accounting for trends prior to the SDIL announcement there were absolute reductions in daily consumption of free sugars from the whole diet in children and adults of 4.8g(95%CI: 0.6g to 9.1g) and 10.9g(95%CI: 7.8g to 13.9g), respectively. Comparable reductions in free sugar consumption from drinks alone were 3.0g(95%CI: 0.1g to 5.8g) and 5.5g(95%CI: 2.7g to 8.3g). The percentage of total dietary energy from free sugars declined over the study period but wasn’t significantly different to the counterfactual.

**Conclusion:** The SDIL led to significant reductions in dietary free sugar consumption in children and adults. Energy from free sugar as a percentage of total energy did not change relative to the counterfactual which could be due to simultaneous reductions in total energy intake associated with reductions in dietary free sugar.

**WHAT IS ALREADY KNOWN ON THIS TOPIC:** High intakes of free sugars are associated with a range of non-communicable diseases. Sugar sweetened beverages constitute a major source of dietary free sugars in children and adults.

The UK Soft Drink Industry levy (SDIL) led to a reduction in the sugar content in many sugar sweetened beverages; and a reduction in household purchasing of sugar from drinks.

No previous study has examined impacts of the SDIL on total dietary consumption of free sugars at the individual level

**WHAT THIS STUDY ADDS:** There were declining trends in intake of dietary free sugar in adults and children prior to the UK SDIL

Accounting for prior trends, one year after the UK SDIL came into force, children and adults further reduced their free sugar intake from food and drink by ∼5g and 11g/day, respectively. Children and adults reduced their daily free sugar intake from soft drinks alone by ∼3g and ∼6g/day, respectively.

Energy intake from free sugars as a proportion of total energy consumed did not change significantly following the UK SDIL, indicating energy intake from free sugar was reducing simultaneously with overall total energy intake.

**HOW THIS STUDY MIGHT AFFECT RESEARCH, PRACTICE OR POLICY:** The UK SDIL was associated with significant reductions in consumption of free sugars from soft drinks and across the whole diet and reinforces previous research indicating a reduction in purchasing. This evidence should be used to inform policy when extending or considering other sugar reduction strategies.

Energy intake from free sugars has been falling but levels remain higher than the 5% recommendation set by the World Health Organization. Reductions in dietary sugar in relation to the SDIL may have driven significant reductions in overall energy.

## Introduction

High consumption of free sugars is associated with non-communicable diseases^1^ including dental caries^2^, obesity^3^, heart disease^4^ and diabetes^1^. Guidelines from the World Health Organization (WHO) and the UK Scientific Advisory Committee on Nutrition (SACN) suggest limiting free sugar consumption^1^ to below 5% of total energy intake to achieve maximum health benefits^1,5^ equivalent to daily maximum amounts of 30g for adults, 24g for children (7-10 years) and 19g for young children (4-6 years). In the United Kingdom (UK) consumption of free sugar is well above the recommended daily maximum, although levels have fallen over the last decade^6^. For example, adolescents consume approximately 70 grams/day^7^ and obtain 12.3% of their energy from free sugars^6^. Sugar sweetened beverages constitute a major source of free sugar in the UK diet^8^,^5^. A growing body of evidence has demonstrated a link between consumption of SSBs and higher risk of weight gain, type II diabetes, coronary heart disease, and premature mortality^9^ such that the WHO recommends taxation of SSBs in order to reduce over-consumption of free sugars and to improve health ^10^. To date, >50 countries have introduced taxation on SSBs which has been associated with a reduction in sales and dietary-intake of free sugar from SSBs^11^. Reductions in childhood obesity prevalence ^12,13^ and improvements in dental health outcomes^14–16^ have also been reported.

In March 2016, the UK government announced the UK soft drink industry levy (SDIL), a two-tier levy on manufacturers, importers and bottlers of soft drinks, would come into force in March 2018^17^. The levy was designed to incentivise manufacturers to reformulate and reduce the free sugar content of SSBs. Details can be found in online supplemental text 1.

One year after the UK SDIL was implemented, there was evidence for a reduction in the sugar content of soft drinks^18^ and households, on average reduced the amount of sugar purchased from soft drinks by 8g/week with no evidence of substitution to confectionary or alcohol. ^19^ However, lack of available data meant it was not possible to examine substitution of purchasing to other sugary foods and drinks, which has previously been suggested in some but not all studies^20,21^. Household purchasing only approximates individual consumption, because it captures only those products brought into the home, products may be shared unequally between household members, and does not account for waste.

To examine effects of the SDIL on total sugar intake at the individual level, in this study we used surveillance data collected using 3 or 4 day-food diaries as part of the UK National Diet and Nutrition Survey (NDNS). We aimed to examine changes in absolute and relative consumption of free sugars from soft drinks alone and from both food and drinks (allowing us to consider possible substitutions to other sugary food items), following the announcement and implementation of the UK SDIL.

## Methods

### Data source

We used eleven years of data (2008 -2019) from the NDNS. Data collection, sampling design and information on response is described in full elsewhere^22^. In brief, NDNS is a continuous, national cross-sectional survey capturing information on food consumption, nutritional status, and nutrient intake inside and outside of the home, in a representative annual sample of ∼ 500 adults and 500 children (1.5-18 years) living in private households in the UK. Participants are sampled throughout the year, such that in a typical month ∼ 40 adults and 40 children participate. For further details see online supplementary text 2

### Outcomes of interest

Outcomes of interest were absolute and relative changes in the total intake of dietary free sugar from i) all food and drinks combined and ii) from non-alcoholic drinks alone. A definition of free sugar can be found in online supplemental text 3. Drink categories examined were those that fell within the following NDNS categories: soft drinks -not low calorie; soft drinks -low calorie; semi-skimmed milk; whole milk; skimmed milk; fruit juice, 1% fat milk and other milk and cream. Additionally, we examined absolute and relative changes in % energy from free sugar in i) food and drinks and ii) drinks alone. While examination of changes in sugar consumption and % energy from sugar across the whole diet (food and drink) captures overall substitutions to other sugar-containing products following the UK SDIL, examination of sugar consumption from drinks alone provides a higher level of specificity to the SDIL.

Protein intake was selected as a non-equivalent dependant control; it was not a nutritional component specifically targeted by the intervention or other government interventions and therefore unlikely to be affected by the SDIL but could still be affected by confounding factors such as increases in food prices^23^ (See online supplementary text 4).

### Statistical analysis

Controlled Interrupted time series (CITS) analyses were performed to examine changes in the outcomes in relation to the UK SDIL separately in adults and children. We analysed data at the quarterly level over eleven years from April 2008 to January 2019. Where diary date entries extended over two quarters, the earlier quarter was designated as the time point for analysis. Generalised least squares (GLS) models were used. Autocorrelation in the time series was determined using Durbin-Watson tests and from visualisations of autocorrelation and partial correlation plots. Autocorrelation-moving average correlation structure with order (p) and moving average (q) parameters were used and selected to minimise the Akaike information criterion (AIC) in each model. Trends in free sugar consumption prior to the announcement of SDIL in April 2016 were used to estimate counterfactual scenarios of what would have happened if the SDIL had not been announced or come into force. Thus, the interruption point was the three-month period beginning April 2016. Absolute and relative differences in consumption of free sugars/person/day were estimated by calculating the difference between the observed and counterfactual values at quarterly time point 45 (Jan-March 2019). To account for non-response, weights provided by NDNS were used and adapted for analysis of adults and children separately^24^.A study protocol has been published^25^ and the study is registered (ISRCTN18042742). For changes to the original protocol see online supplemental text 5.

## Results

Data from 7999 adults and 7656 children were included across 11 years representing ∼40 children and ∼40 adults each month. Table 1 gives descriptive values for the outcomes of interest. In the post-announcement period compared to the pre-announcement period, free sugars consumed from all drinks reduced by around a half in children and a third in adults. Total dietary free sugar consumption and percentage of total dietary energy derived from free sugars also declined. Mean protein consumption was relatively stable over both periods in children and adults. The age and sex of children and adults were very similar in the pre and post-announcement period.

**Table 1:**
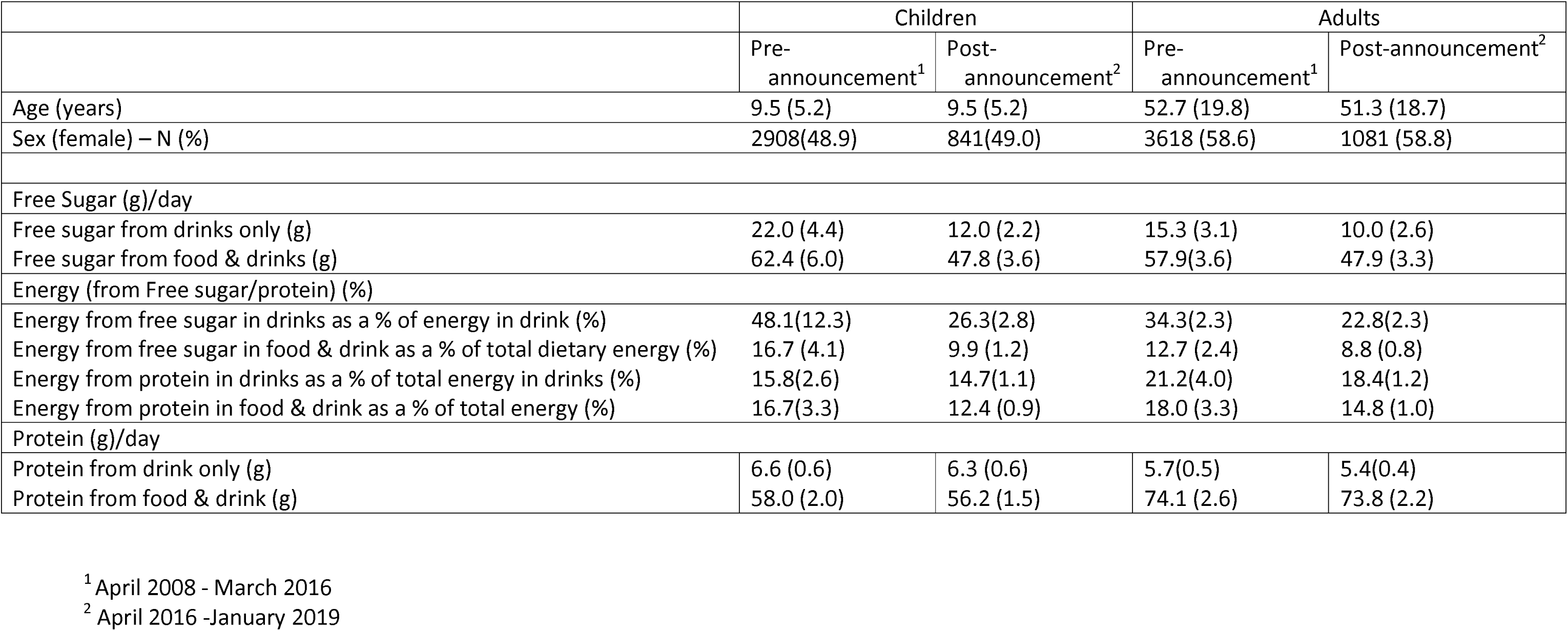
Mean amount of free sugar (g) consumed in children or adults per day during the study period before and after the announcement of SDIL.

|  | Children |  | Adults |  |
| --- | --- | --- | --- | --- |
|  | Pre-announcement<sup>1</sup> | Post-announcement<sup>2</sup> | Pre-announcement<sup>1</sup> | Post-announcement<sup>2</sup> |
| Age (years) | 9.5 (5.2) | 9.5 (5.2) | 52.7 (19.8) | 51.3 (18.7) |
| Sex (female) – N (%) | 2908(48.9) | 841(49.0) | 3618 (58.6) | 1081 (58.8) |
| Free Sugar (g)/day |  |  |  |  |
| Free sugar from drinks only (g) | 22.0 (4.4) | 12.0 (2.2) | 15.3 (3.1) | 10.0 (2.6) |
| Free sugar from food & drinks (g) | 62.4 (6.0) | 47.8 (3.6) | 57.9(3.6) | 47.9 (3.3) |
| Energy (from Free sugar/protein) (%) |  |  |  |  |
| Energy from free sugar in drinks as a % of energy in drink (%) | 48.1(12.3) | 26.3(2.8) | 34.3(2.3) | 22.8(2.3) |
| Energy from free sugar in food & drink as a % of total dietary energy (%) | 16.7 (4.1) | 9.9 (1.2) | 12.7 (2.4) | 8.8 (0.8) |
| Energy from protein in drinks as a % of total energy in drinks (%) | 15.8(2.6) | 14.7(1.1) | 21.2(4.0) | 18.4(1.2) |
| Energy from protein in food & drink as a % of total energy (%) | 16.7(3.3) | 12.4 (0.9) | 18.0 (3.3) | 14.8 (1.0) |
| Protein (g)/day |  |  |  |  |
| Protein from drink only (g) | 6.6 (0.6) | 6.3 (0.6) | 5.7(0.5) | 5.4(0.4) |
| Protein from food & drink (g) | 58.0 (2.0) | 56.2 (1.5) | 74.1 (2.6) | 73.8 (2.2) |

<sup>1</sup> April 2008 - March 2016
<sup>2</sup> April 2016 -January 2019

All estimates of change in free sugar consumption referred to below are based on grams/individual/day, in the three-month period beginning January 2019 and compared to the counterfactual scenario of no UK SDIL announcement or implementation.

### Change in free sugar consumption (drinks only)

In children, consumption of free sugars from drinks was ∼ 27g/day at the start of the study period but fell steeply throughout. By the end of the study period mean sugar consumption from drinks was ∼ 10g/day (Figure 1). Overall, there was an absolute reduction in daily free sugar consumption from drinks of 3.0g (95% CI 0.1 to 5.8) or a relative reduction of 23.5% (95% CI 46.0% to 0.9%) in children (Table 2). In adults, free sugar consumption at the beginning of the study was lower than that of children (∼17g/day) and was declining prior to the SDIL announcement, albeit less steeply (Figure 1). Following the SDIL announcement, free sugar consumption from drinks appeared to decline even more steeply. There was an absolute reduction in free sugar consumption from drinks of 5.2g (95% CI 4.2g to 6.1g) or a relative reduction of 40.4% (95% CI 32.9% to 48.0%) in adults (Figure 1, Table 2).

**Figure 1:**
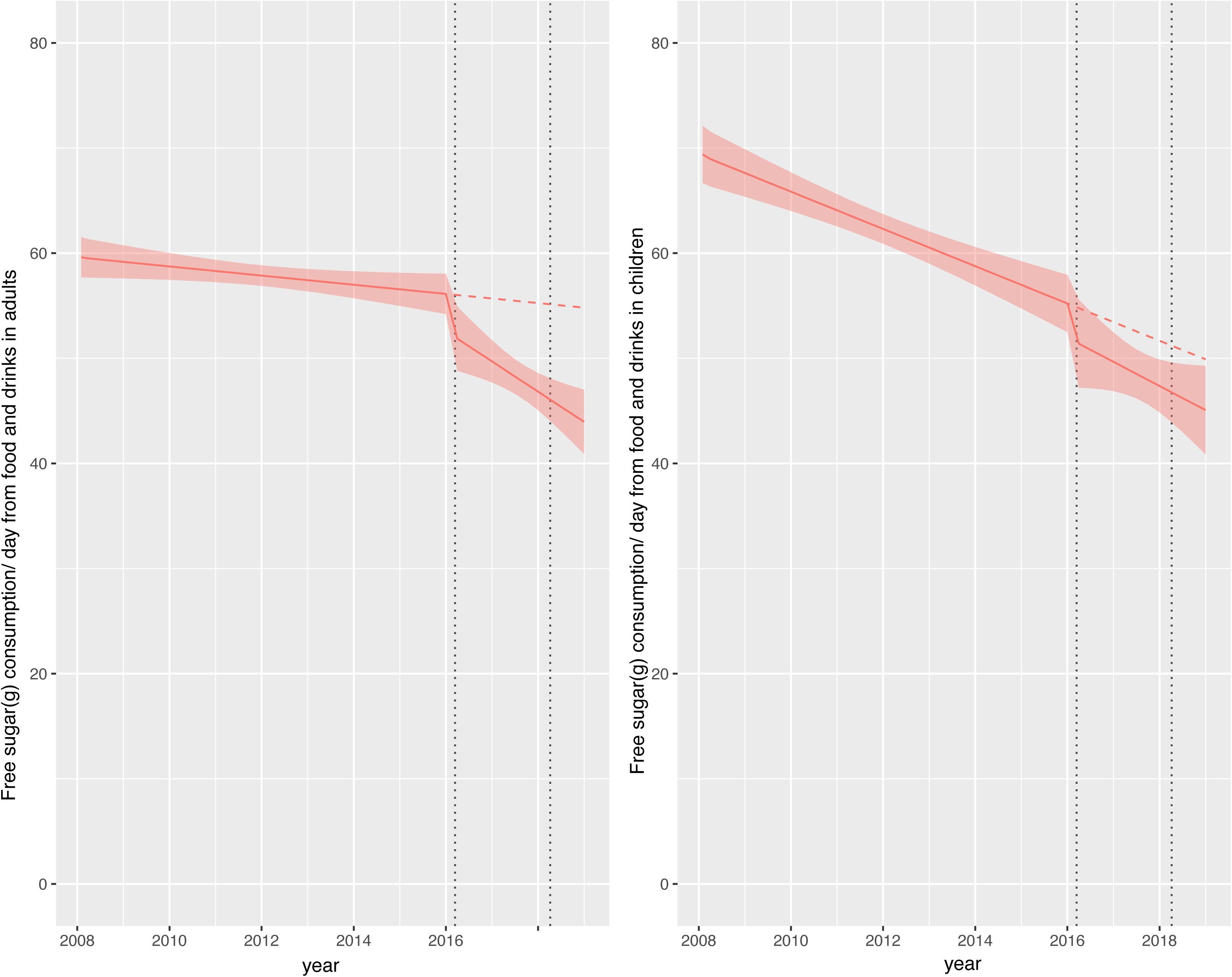
Observed and modelled daily consumption (g) of free-sugar from drink products per adult/children from January 2008 to February 2020. Red points show observed data and solid red lines (with light red shadows) shows modelled data (and 95% confidence intervals) of free-sugar consumed from drinks. The dashed red line indicates the counterfactual line based on preannouncement trends and had the announcement and implementation not happened. Modelled protein consumption from drinks (control group) was removed from the graph to include resolution but is available in the supplementary section. The first and second dashed lines indicate the announcement and implementation of SDIL, respectively.

**Table 2:** Change in free sugar consumption in food and drink and energy from free sugar as a proportion of total energy compared to the counterfactual scenario of no announcement and implementation of the UK SDIL

|  | Children |  | Adults |  |
| --- | --- | --- | --- | --- |
|  | Absolute change (g) | Relative change (%) | Absolute change (g) | Relative change (%) |
| Free sugar from drinks only | **-3.0(-5.8, -0.1)** | **-23.5 (-46.0, -0.9)** | **-5.2 (-6.1, -4.2)** | **-40.4 (-48.0, -32.9)** |
| Free sugar from food and drinks | **-4.8 (-9.1, -0.6)** | **-9.7( -18.2, -1.2)** | **-10.9 (-13.9, -7.8)** | **-19.8 (-25.4, -14.2)** |
| energy from free sugar in food & drink as a % of total energy (%) | -0.7 (-3.9, 2.5) | -7.6 (-41.7, 26.5) | -2.6 (0.6, -5.8) | -24.3 (-54.0,5.4) |
| Energy from free sugar in drink as a % of total energy in drink (%) | 0.4 (-7.1, 8.0) | 1.8 (-30.7, 34.3) | -0.52(-5.4, 4.3) | -2.4 ( -24.6,19.8) |

### Change in total dietary free sugar consumption (food and drinks combined)

Consumption of total dietary free sugars in children was ∼ 70g/day at the beginning of the study but this fell to ∼ 45g/day by the end of the study (Figure 2). Relative to the counterfactual scenario, there was an absolute reduction in total dietary free sugar consumption of 4.8g (95% CI 0.6 to 9.1) or relative reduction of 9.7% (95% CI 18.2% to 1.2%) in children (Figure 2; Table 2). In adults, consumption of total dietary free sugar consumption at the beginning of the study was ∼60g/day falling to ∼45g/day by the end of the study (Figure 2). Relative to the counterfactual scenario there was an absolute reduction in total dietary free sugar consumption in adults of 10.9g (95% CI 7.8g to 13.9g) or a relative reduction of 19.8% (95% CI 25.4% to 14.2%). Supplementary figures show that dietary protein consumption and energy from protein was more-or-less stable across the study period (supplementary figures S1-S4).

**Figure 2:**
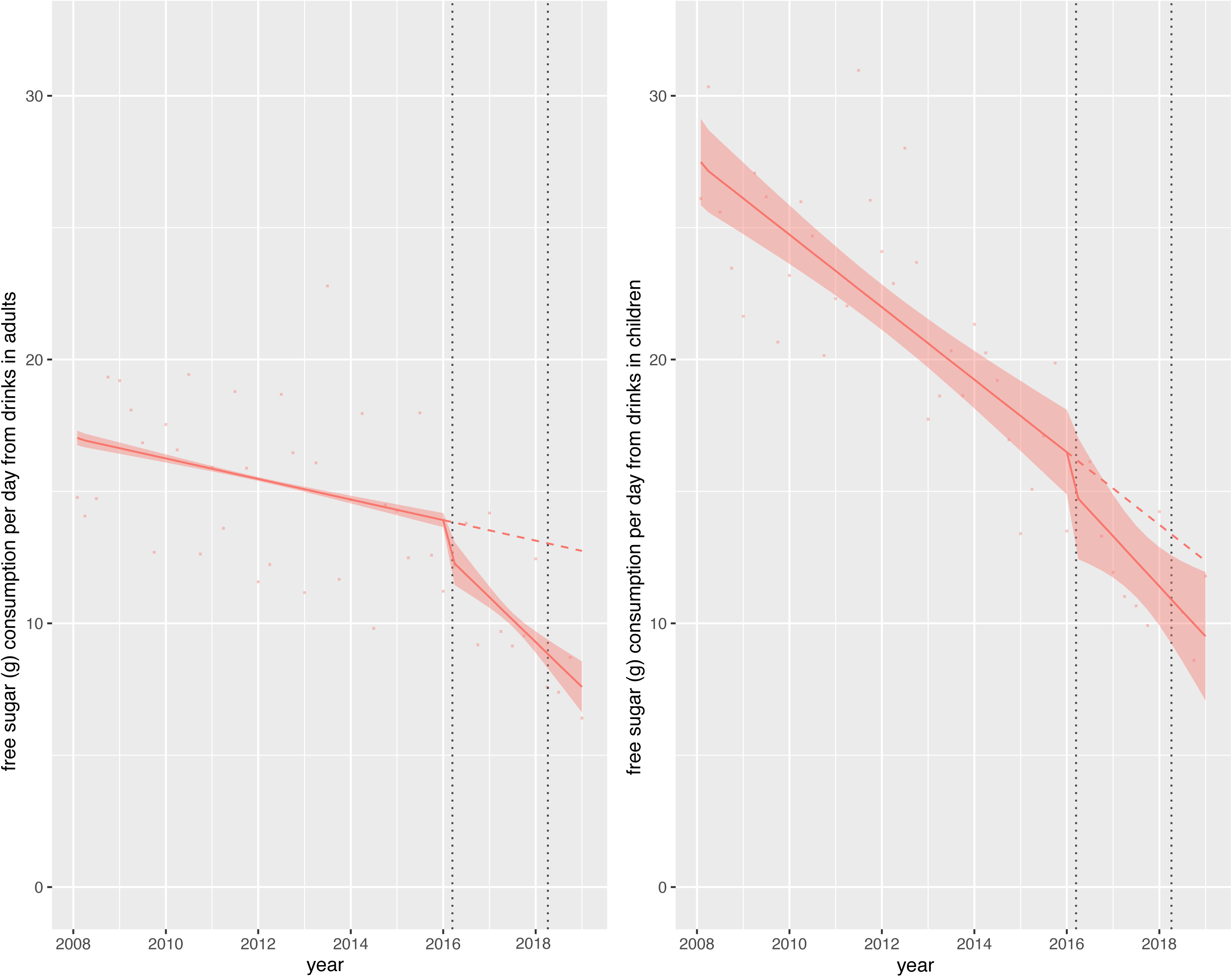
Observed and modelled daily consumption (g) of free-sugar from food and drink products per adult/children from January 2008 to February 2020. Red points show observed data and solid red lines (with light red shadows) shows modelled data (and 95% confidence intervals) of free-sugar consumed from food and drinks. The dashed red line indicates the counterfactual line based on preannouncement trends and had the announcement and implementation not happened. Modelled protein consumption from food and drinks (control group) was removed from the graph to include resolution but is available in the supplementary section. The first and second dashed lines indicate the announcement and implementation of SDIL, respectively.

### Change in energy from free sugar as a proportion of total energy

The % of energy from total dietary free sugar decreased across the study period but did not change significantly relative to the counterfactual scenario, in children or adults, with relative changes in free sugar consumption of -7.6g (95% CI -41.7 to 26.5) and -24.3(95% CI -54.0 to 5.4), respectively (Figure 3, Table 2). Energy from free sugar in drinks as a proportion of total energy from drinks also decreased across the study period but did not change significantly relative to the counterfactual (Figure 4).

**Figure 3:**
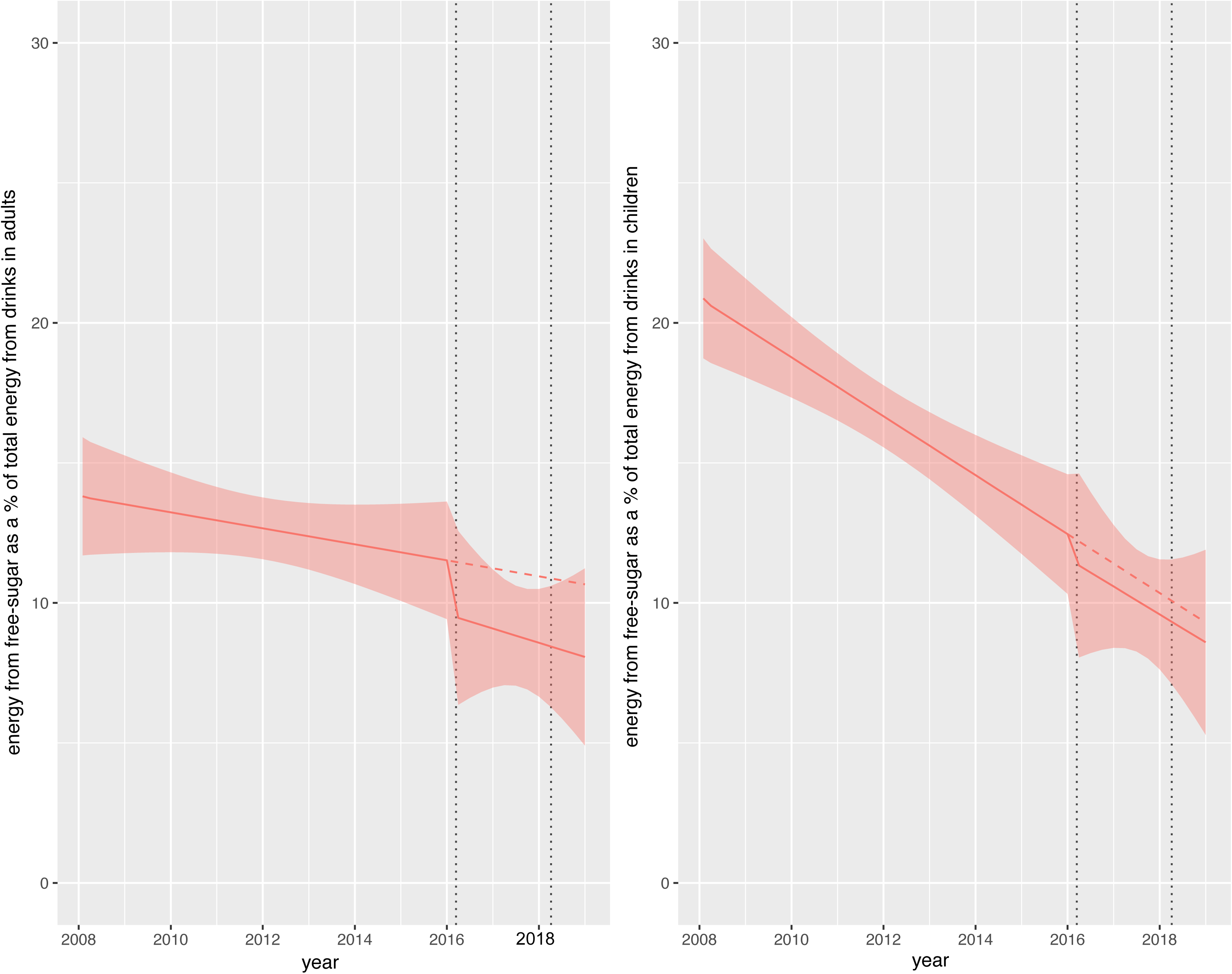
Observed and modelled energy from free-sugar in food and drinks as a % of energy from food and drinks in adult/children from January 2008 to February 2020. Red points show observed data and solid red lines (with light red shadows) shows modelled data (and 95% confidence intervals) of energy from free-sugar in drinks as a % of total energy intake. The dashed red line indicates the counterfactual line based on preannouncement trends and had the announcement and implementation not happened. Modelled energy from protein consumption in food and drinks (control group) was removed from the graph to include resolution but is available in the supplementary section. The first and second dashed lines indicate the announcement and implementation of SDIL, respectively.

**Figure 4:**
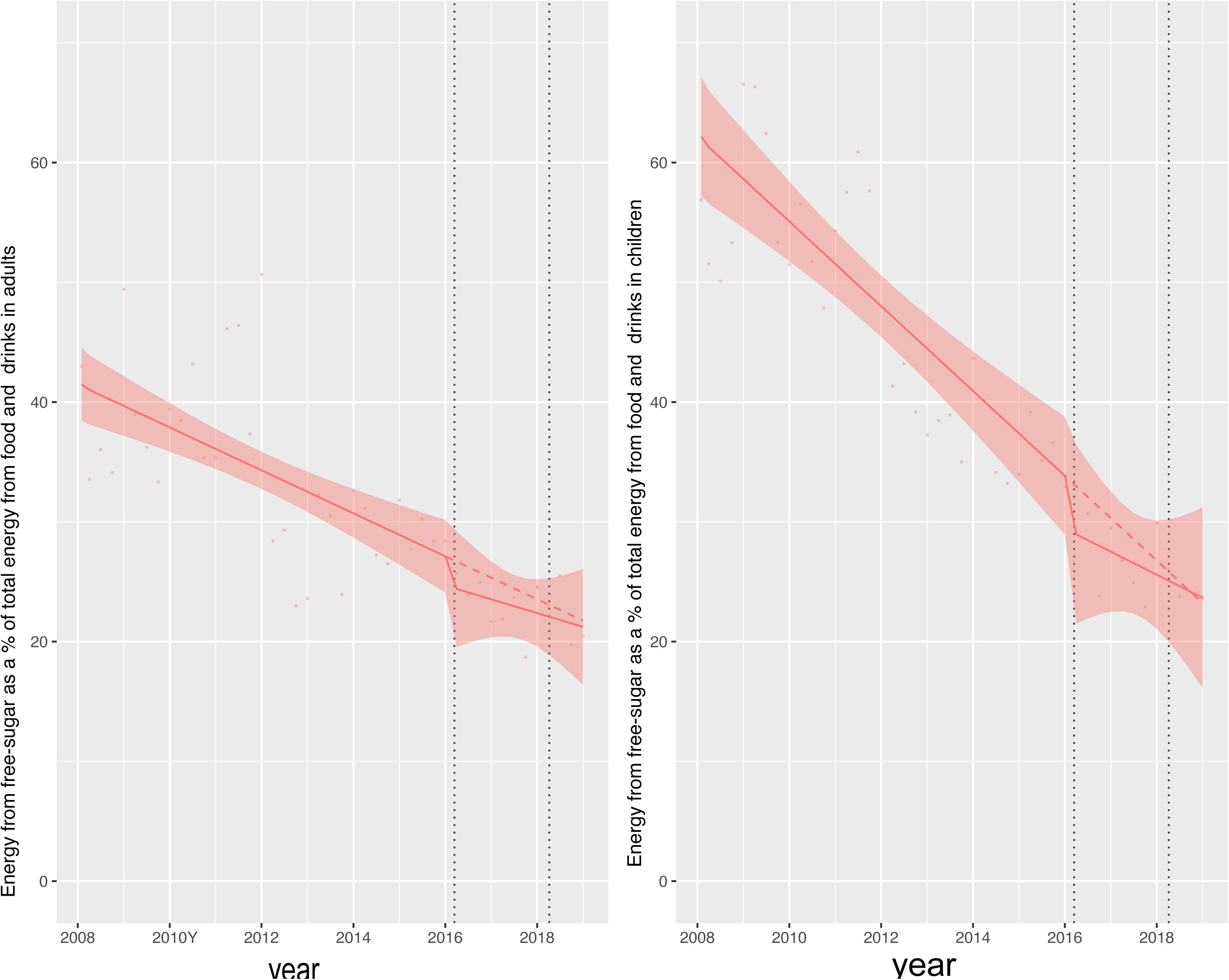
Observed and modelled energy from free-sugar in drinks as a % of energy from drinks in adult/children from January 2008 to February 2020. Red points show observed data and solid red lines (with light red shadows) shows modelled data (and 95% confidence intervals) of energy from free-sugar in drinks as a % of total energy intake. The dashed red line indicates the counterfactual line based on preannouncement trends and had the announcement and implementation not happened. Modelled energy from protein consumption in drinks (control group) was removed from the graph to include resolution but is available in the supplementary section. The first and second dashed lines indicate the announcement and implementation of SDIL, respectively.

## Discussion

### Summary of main findings

This study is the first to examine individual level consumption of free sugars in the total diet (and in drinks only) in relation to the UK SDIL. Using nationally representative population samples we found that approximately one year following the UK SDIL, there was a reduction in total dietary free sugar consumed by children and adults compared to what would have been expected if the SDIL had not been announced or implemented. In children, this was equivalent to a reduction of 4.8g of free sugars/day from food and drinks, of which 3g/day came from drinks alone, suggesting that the reduction of sugar in the diet was primarily due to a reduction of sugar from drinks. In adults, reductions in dietary sugar appeared to come equally from food and drink with an 11g reduction in food and drink combined of which 5.5g was from drinks only. There was no significant reduction compared to the counterfactual in the percentage of energy intake from free sugars in the total diet or from drinks alone, in both children and adults, suggesting that energy intake from free sugar was reducing simultaneously with overall total energy intake.

### Comparison with other studies and Interpretation of results

Our finding of a reduction in consumption of free sugars from soft drinks after accounting for pre-SDIL announcement trends is supported by previous research showing a large reduction in the proportion of available drinks with over 5g of sugar /100ml, the threshold in which soft drinks become levy liable^18^. Furthermore, efforts of the soft drink industry to reformulate soft drinks were found to have led to significant reductions in the volume and per capita sales of sugar from these drinks^26^.

Our findings are consistent with recent research showing reductions in purchasing of sugar from soft drinks^19^of approximately 8g/household/week (equivalent to ∼3g/person/week or ∼0.5g/ person/day), one year after the SDIL came into force^19^.The estimates from the current study suggest larger reductions in consumption (eg:3g free sugar/ day from drinks in children) than previously reported for purchasing. Methodological differences may explain these differences in estimated effect sizes. Most importantly, the previous study used data on soft drink purchases that were for consumption in the home only. In contrast, here we captured information on consumption (rather than purchasing) in and out of the home. Consumption of food and particularly drinks outside of the home in young people (1-21 years) increases with age^7^ and makes a substantial contribution to total free sugar intakes, highlighting the importance in recording both in and out-of-home sugar consumption ^7^. Purchasing and consumption data also treat waste differently – purchase data records what comes into the home and therefore includes waste, whereas consumption data specifically aims to capture leftovers and waste and exclude it from consumption estimates. While both studies use weights to make the population samples representative of the UK, there may be differences in the study participant characteristics in the two studies, which may contribute to the different estimates.

Consistent with other studies^27^, we found that across the 11 year study period we observed a downward trend in free sugar and energy intake in adults and children ^6^. A decline in consumption of free sugars was observed in the whole diet rather than just drinks suggesting that consumption of free sugar from food was also declining from as early as 2008. One reason might be the steady transition from sugar in the diet to low-calorie artificial sweeteners which globally have had an annual growth of approximately 5.1% between 2008 and 2015^28^

Public health signalling around the time of the announcement of the levy may also have contributed to the changes we observed. Public acceptability and perceived effectiveness of the SDIL was reported to be high four months before and ∼ twenty months after the levy came into force^29^ Furthermore, awareness of the SDIL was found to be high amongst parents of children living in the UK, with most supporting the levy and intending to reduce purchases of SSBs as a result^30^. Health signalling was also found following the implementation of the SSB tax in Mexico, with one studying reporting that most adults (65%) were aware of the tax and those aware of the tax were more likely to think the tax would reduce purchases of SSB ^31^ although a separate study found that adolescents in Mexico were mostly unaware of the tax ^32^ suggesting that public health signalling may differ according to age.

In 2016 the UK government announced a voluntary sugar reduction programme, as part of its childhood obesity plan, with the aim of reducing sugar sold by industry by 20% through both reformulation and portion size reduction^33^. While the programme only managed to achieve overall reductions of ∼3.5% this did include higher reductions in specific products such as yogurts (13.5%) and cereals (14.9%) and may have contributed to the observed reductions in total sugar consumption around the time of the SDIL.

Our findings, consistent with previous analyses^6^, showed there was a downward trend in energy intake from sugar as a proportion of total energy across the duration of the study. While there was no reduction compared to the counterfactual scenario (which was also decreasing), our estimates suggest that by 2019 on average energy from sugar as a proportion of all energy appears to be in line with the WHO recommendation of 10%^1^ but not the more recent guidelines of 5% which may bring additional health benefits^1,34^. This finding may suggest that reductions in energy intake from sugar were reducing in concert with overall energy intake and indeed may have been driving it.

However, the magnitude of calories associated with the reduction in free sugars, compared to the counterfactual scenario in both adults and children, was modest and thus potentially too small to reflect significant changes in % energy from sugar. In children, a daily reduction of 4.8g sugar equates to ∼19.2 kilocalories out of an approximate daily intake of ∼2000 kilocalories which is equivalent to ∼1% reduction in energy intake. Furthermore, overall measures of dietary energy are also likely to involve a degree of error reducing the level of precision in any estimates.

Our estimates of changes in sugar consumption in relation to SDIL suggest that adults may have experienced a greater absolute reduction in sugar than children, which is not consistent with estimates of the distributional impact of the policy^35^. However, our understanding may be aided with the visualisations afforded by graphical depictions of our ITS graphs. Children’s consumption of sugar at the beginning of the study period, particularly in drinks, was higher than in adults but reducing at a steeper trajectory, which will have influenced our estimated counterfactual scenario of what would have happened without the SDIL. This steep downward trajectory could not have continued indefinitely, as there is a lower limit for sugar consumption. No account for this potential ‘floor effect’ was made in the counterfactual. Adults had a lower baseline of sugar consumption, but their trajectory of sugar consumption were decreasing at a gentler trajectory potentially allowing more scope for improvement over the longer run.

Reductions in the levels of sugar in food and drink may have also impacted different age-groups and children and adults, differently. For example, the largest single contributor to free sugars in younger children between the ages of 4-10y is cereal and cereal products, followed by soft drinks and fruit juice. By the age of 11 to 18 years, soft drinks provide the largest single source (29%) of dietary free sugar. For adults the largest source of free sugars is sugar, preserves and confectionery, followed by non-alcoholic beverages^8^.

### Strengths and limitations

The main strengths of the study include the use of nationally representative data on individual consumption of food and drink in and out of the home, using consistent food diary assessment over a four-day period, setting it apart from other surveys which have used food frequency questionnaires, 24 hour recall, shortened dietary instruments or a mixture of these approaches across different survey years^36^. The continual collection of data using consistent methods enabled us to analyse dietary sugar consumption and energy quarterly, over 11 years (or 45 time points) including the announcement and implementation period of the SDIL. Information on participant age allowed us to examine changes in sugar consumption in adults and children separately. Limited sample sizes restricted our use of weekly or monthly data and prevented us from examining differences between sociodemographic groups. At each time point we used protein consumption in food and drink as a non-equivalent control category, strengthening our ability to adjust for time-varying confounders such as contemporaneous events. The trends in counterfactual scenarios of sugar consumption and energy from free sugar as part of total energy were based on trends from April 2008 to the announcement of the UK SDIL (March 2016) however it is possible that the direction of sugar consumption may have changed course. Ascribing changes in free sugar consumption to the SDIL should include exploration of other possible interventions that might have led to a reduction in sugar across the population. We are only aware of the wider UK government’s voluntary sugar reduction programme^37^, implemented across overlapping timelines (2015-2020).

This aimed to cut sugar in food products by 20% by 2020 and was predicted to reduce levels of obesity^38^. The sugar reduction programme was however found to have led to much smaller reductions of ∼3.5% in purchasing of sugar in non-drinks categories^37^. In turn, underreporting of portion sizes and high energy foods, which may be increasingly seen as less socially acceptable has been suggested as a common error in self-reported dietary intake however there is no evidence to suggest this would have changed as a direct result of the SDIL^39^

## Conclusions

Our findings indicate that the UK SDIL led to reductions in consumption of dietary free sugars in adults and children, one year after the SDIL came into force. Energy from free sugar as a proportion of overall energy intake was falling prior to the UK SDIL but did not change in relation to the SDIL suggesting that a reduction in sugar may have driven a simultaneous reduction in overall energy intake.

## Ethics Statement

NDNS participants provided written informed consent to take part in the study. Analysis of this anonymised data did not require prior ethical approval and is obtainable to eligible researchers from the UK Data Archive. Ethical approval for NDNS was approved from the Oxfordshire A Research Ethics Committee. All statistical analyses were performed in R version 4.1.0.

## Funding

NTR, OM, MW, and JA were supported by the Medical Research Council (grant Nos MC_UU_00006/7). This project was funded by the NIHR Public Health Research programme (grant Nos 16/49/01 and 16/130/01) to MW. The views expressed are those of the authors and not necessarily those of the National Health Service, the NIHR, or the Department of Health and Social Care, UK. The funders had no role in study design, data collection and analysis, decision to publish, or preparation of the manuscript.

## Competing interests

The authors of this manuscript have no competing interests.

## Data Availability

NDNS data are made publicly available via the UK Data Service (UKDS).

https://ukdataservice.ac.uk/

## Supplementary Figures

**Figure S1:** Observed and modelled daily consumption (g) of protein from drink products per adult/children from January 2008 to February 2020. Red points and solid red lines (with 95% CI) show modelled protein consumption from drinks. The dashed red line indicates the counterfactual line based on preannouncement trends in protein consumption and had the announcement and implementation not happened. The light blue lines represents free-sugar consumption (g) from drinks as shown in figure 1. The first and second dashed lines indicate the announcement and implementation of SDIL, respectively.

**Figure S2:** Observed and modelled daily consumption (g) of protein from food and drink products per adult/children from January 2008 to February 2020. Red points and solid red lines (with 95% CI) show modelled protein consumption from food and drinks. The dashed red line indicates the counterfactual line based on preannouncement trends in protein consumption and had the announcement and implementation not happened. The light blue lines represent free-sugar consumption (g) from food and drinks as shown in figure 2. The first and second dashed lines indicate the announcement and implementation of SDIL, respectively.

**Figure S3:** Observed and modelled energy from protein as a % of energy from drinks in adult/children from January 2008 to February 2020. Red points show observed data and solid red lines (with light red shadows) shows modelled data (and 95% confidence intervals) of energy from protein in drinks as a % of total energy intake. The dashed red line indicates the counterfactual line based on preannouncement trends and had the announcement and implementation not happened. The light blue lines represent energy from free-sugar from drinks, as shown in figure 4. The first and second dashed lines indicate the announcement and implementation of SDIL, respectively.

**Figure S4:** Observed and modelled energy from protein in food and drinks as a % of energy from food and drinks in adult/children from January 2008 to February 2020. Red points show observed data and solid red lines (with light red shadows) shows modelled data (and 95% confidence intervals) of energy from protein in food and drinks as a % of total energy intake. The dashed red line indicates the counterfactual line based on preannouncement trends and had the announcement and implementation not happened. The light blue lines represent energy from free-sugar from food and drinks, as shown in figure 3. The first and second dashed lines indicate the announcement and implementation of SDIL, respectively.

## Notes

### Competing Interest Statement

The authors have declared no competing interest.

### Author Declarations

NDNS participants provided written informed consent to take part in the study. Analysis of this anonymised data did not require prior ethical approval and is obtainable to eligible researchers from the UK Data Archive. Ethical approval for NDNS was approved from the Oxfordshire A Research Ethics Committee.

## References

1. World Health Organization. Guideline: Sugars intake for adults and children. 2015.

2. Moynihan P, Kelly S. Effect on caries of restricting sugars intake: systematic review to inform WHO guidelines. J Dent Res 2014; 93: 8–18.

3. Morenga L Te, Mallard S, Mann J. Dietary sugars and body weight: systematic review and meta-analyses of randomised controlled trials and cohort studies. BMJ 2013; 346. DOI:10.1136/BMJ.E7492.

4. Kelly RK, Tong TYN, Watling CZ, et al. Associations between types and sources of dietary carbohydrates and cardiovascular disease risk: a prospective cohort study of UK Biobank participants. BMC Med 2023; 21: 34.

5. Scientific Advisory Committee on Nutrition. Carbohydrates and Health. 2015.

6. National Diet and Nutrition Survey Rolling programme Years 9 to 11 (2016/2017 to 2018/2019). 2020.

7. Griffith R, O’Connell M, Smith K, Stroud R. What’s on the Menu? Policies to Reduce Young People’s Sugar Consumption. Fisc Stud 2020; 41: 165–97.

8. Roberts C, Steer T, Maplethorpe N, et al. National Diet and Nutrition Survey Results from Years 7 and 8 (Combined) of the Rolling Programme (2014–2015 to 2015–2016). London Public Heal England 2018.

9. Malik VS, Hu FB. The role of sugar-sweetened beverages in the global epidemics of obesity and chronic diseases. Nat Rev Endocrinol 2022; 18: 205–18.

10. World Health Organization. Together Let’s Beat NCDs. Taxes on sugary drinks: Why do it? 2017.

11. Teng A, Jones A, Mizdrak A, Signal L, Genç M, Wilson N. Impact of sugar-sweetened beverage taxes on purchases and dietary intake: Systematic review and meta-analysis. Obes Rev 2019; 20: 1187–204.

12. Rogers NT, Cummins S, Forde H, et al. Associations between trajectories of obesity prevalence in English primary school children and the UK soft drinks industry levy: An interrupted time series analysis of surveillance data. PLOS Med 2023; 20: e1004160.

13. Gračner T, Marquez-Padilla F, Hernandez-Cortes D. Changes in Weight-Related Outcomes Among Adolescents Following Consumer Price Increases of Taxed Sugar-Sweetened Beverages. JAMA Pediatr 2022; 176: 150–8.

14. Rogers N, Conway D, Mytton O, et al. Trends in childhood hospital admissions for carious tooth extractions in England in relation to the UK soft drinks industry levy: an interrupted time series analysis of Hospital Episode Statistics. medRxiv 2023. DOI:doi.org/10.1101/2023.02.27.23286504.

15. Petimar J, Gibson L., Wolff M., et al. Changes in Dental Outcomes After Implementation of the Philadelphia Beverage Tax. Am J Prev Med 2023.

16. Hernández-F M, Cantoral A, Colchero MA. Taxes to Unhealthy Food and Beverages and Oral Health in Mexico: An Observational Study. Caries Res 2021; 55: 183–92.

17. Soft Drinks Industry Levy comes into effect - GOV.UK. 2018.

18. Scarborough P, Adhikari V, Harrington RA, et al. Impact of the announcement and implementation of the UK Soft Drinks Industry Levy on sugar content, price, product size and number of available soft drinks in the UK, 2015-19: A controlled interrupted time series analysis. PLOS Med 2020; 17: e1003025.

19. Pell D, Mytton O, Penney TL, et al. Changes in soft drinks purchased by British households associated with the UK soft drinks industry levy: controlled interrupted time series analysis. BMJ 2021; 372. DOI:10.1136/BMJ.N254.

20. Powell LM, Chriqui JF, Khan T, Wada R, Chaloupka FJ. Assessing the Potential Effectiveness of Food and Beverage Taxes and Subsidies for Improving Public Health: A Systematic Review of Prices, Demand and Body Weight Outcomes. Obes Rev 2013; 14: 110.

21. Gibson L, Lawman H, Bleich S, et al. No Evidence of Food or Alcohol Substitution in Response to a Sweetened Beverage Tax. Am J Prev Med 2021; 60.

22. Venables MC, Roberts C, Nicholson S, et al. Data Resource Profile: United Kingdom National Diet and Nutrition Survey Rolling Programme (2008-19). Int J Epidemiol 2022; 51: E143–55.

23. Lopez Bernal J, Cummins S, Gasparrini A. The use of controls in interrupted time series studies of public health interventions. Int J Epidemiol 2018; 47: 2082–93.

24. Public Health England. NDNS: Time Trend and Income Analyses for Years 1 to 9 (2019). https://www.gov.uk/government/statistics/ndns-time-trend-and-income-analyses-for-years-1-to-9.

25. White M, Scarborough P, Briggs A, et al. Evaluation of the health impacts of the UK Treasury Soft Drinks Industry Levy (SDIL). NIHR Public Heal. Res. Program. 2017.

26. Bandy LK, Scarborough P, Harrington RA, Rayner M, Jebb SA. Reductions in sugar sales from soft drinks in the UK from 2015 to 2018. BMC Med 2020; 18: 1–10.

27. Marriott BP, Hunt KJ, Malek AM, Newman JC. Trends in intake of energy and total sugar from sugar-sweetened beverages in the United States among children and adults, NHANES 2003-2016. Nutrients 2019; 11. DOI:10.3390/nu11092004.

28. Sylvetsky A, Rother K. Trends in the consumption of low-calorie sweeteners. Physiol Behav 2016; 164. Part: 446–50.

29. Adams J, Pell D, Penney TL, Hammond D, Vanderlee L, White M. Public acceptability of the UK Soft Drinks Industry Levy: Repeat cross-sectional analysis of the International Food Policy Study (2017-2019). BMJ Open 2021; 11: 1–11.

30. Gillison F, Grey E, Griffin T. Parents’ Perceptions and Responses to the UK Soft Drinks Industry Levy. J Nutr Educ Behav 2020; 52: 626–31.

31. Lvarez-Sá Nchez CA, Contento I, Jimé Nez-Aguilar A, et al. Does the Mexican sugar-sweetened beverage tax have a signaling effect? ENSANUT 2016. 2018. DOI:10.1371/journal.pone.0199337.

32. Ortega-Avila AG, Papadaki A, Jago R. Exploring perceptions of the Mexican sugar-sweetened beverage tax among adolescents in north-west Mexico: a qualitative study. Public Health Nutr 2018; 21: 618–26.

33. Sugar reduction – industry progress 2015 to 2020. Final report for foods included in the programme and the latest data for drinks included in the Soft Drinks Industry Levy and juices and milk based drinks. Office for Health Improvement and Disparities. 2022.

34. Public Health England. Why 5%? An explanation of SACN’s recommendations about sugars and health. 2015.

35. Briggs ADM, Mytton OT, Kehlbacher A, et al. Health impact assessment of the UK soft drinks industry levy: a comparative risk assessment modelling study. Lancet Public Heal 2017; 2: e15–22.

36. Campbell M, Smith D, Baird J, Vogel C, Moon EG. A critical review of diet-related surveys in England, 1970-2018. Arch Public Heal 2020; 78: 1–18.

37. Public Health England. Sugar reduction: Report on progress between 2015 and 2018. 2019.

38. Amies-Cull B, Briggs ADM, Scarborough P. Estimating the potential impact of the UK government’s sugar reduction programme on child and adult health: modelling study. BMJ 2019; 365. DOI:10.1136/BMJ.L1417.

39. Lennox A, Bluck L, Page P, et al. Appendix X: Misreporting in the National Diet and Nutrition Survey Rolling Programme (NDNS RP): Summary of Results and Their Interpretation. 2014.

