## Supplementary figures and images for "Estimated changes in free sugar consumption one year after the UK Soft drinks industry levy came into force: controlled interrupted time series analysis of the National Diet and Nutrition Survey (2011-2019)"

### Supplementary fig 1

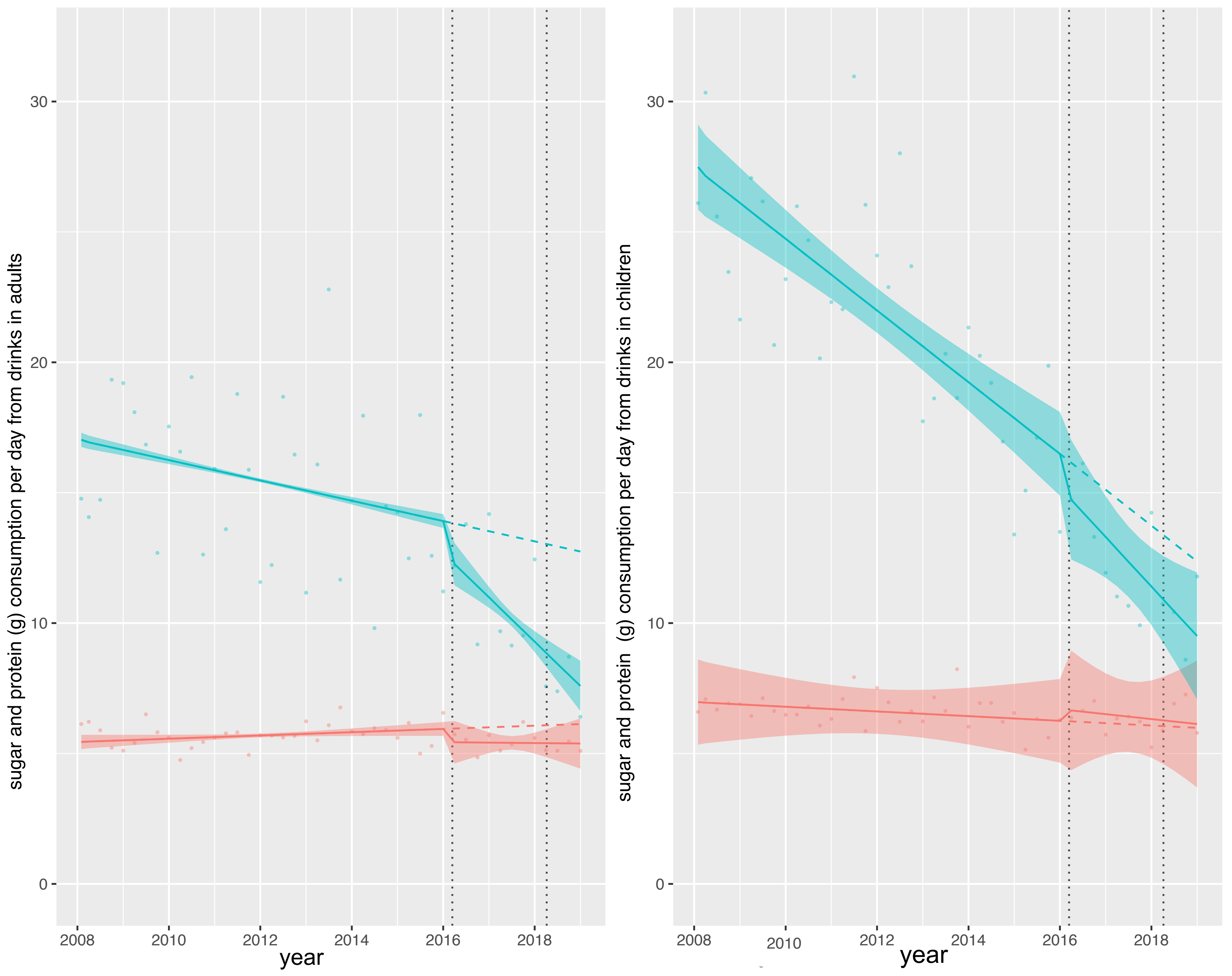

### Supplementary fig 2

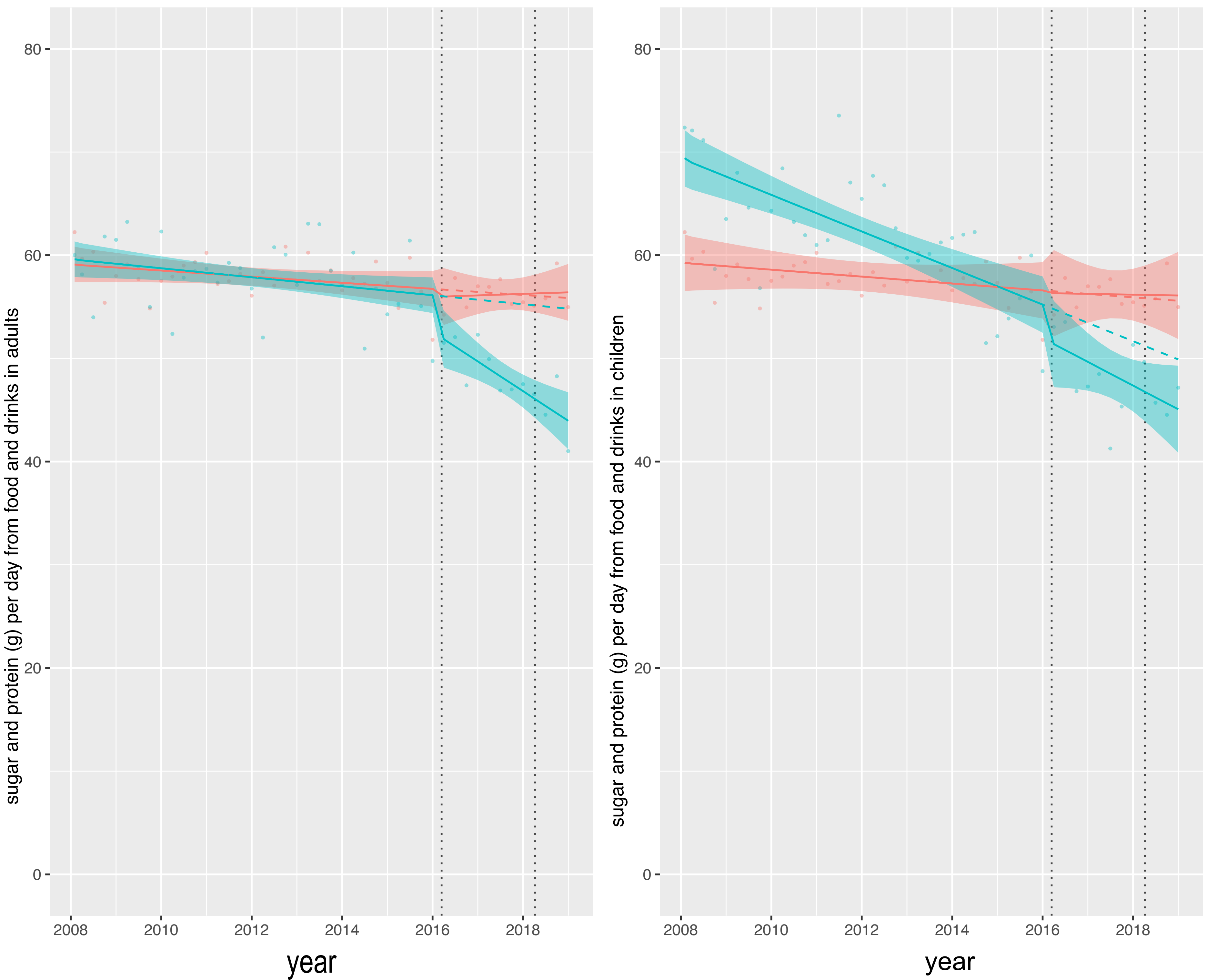

### Supplementary fig 3

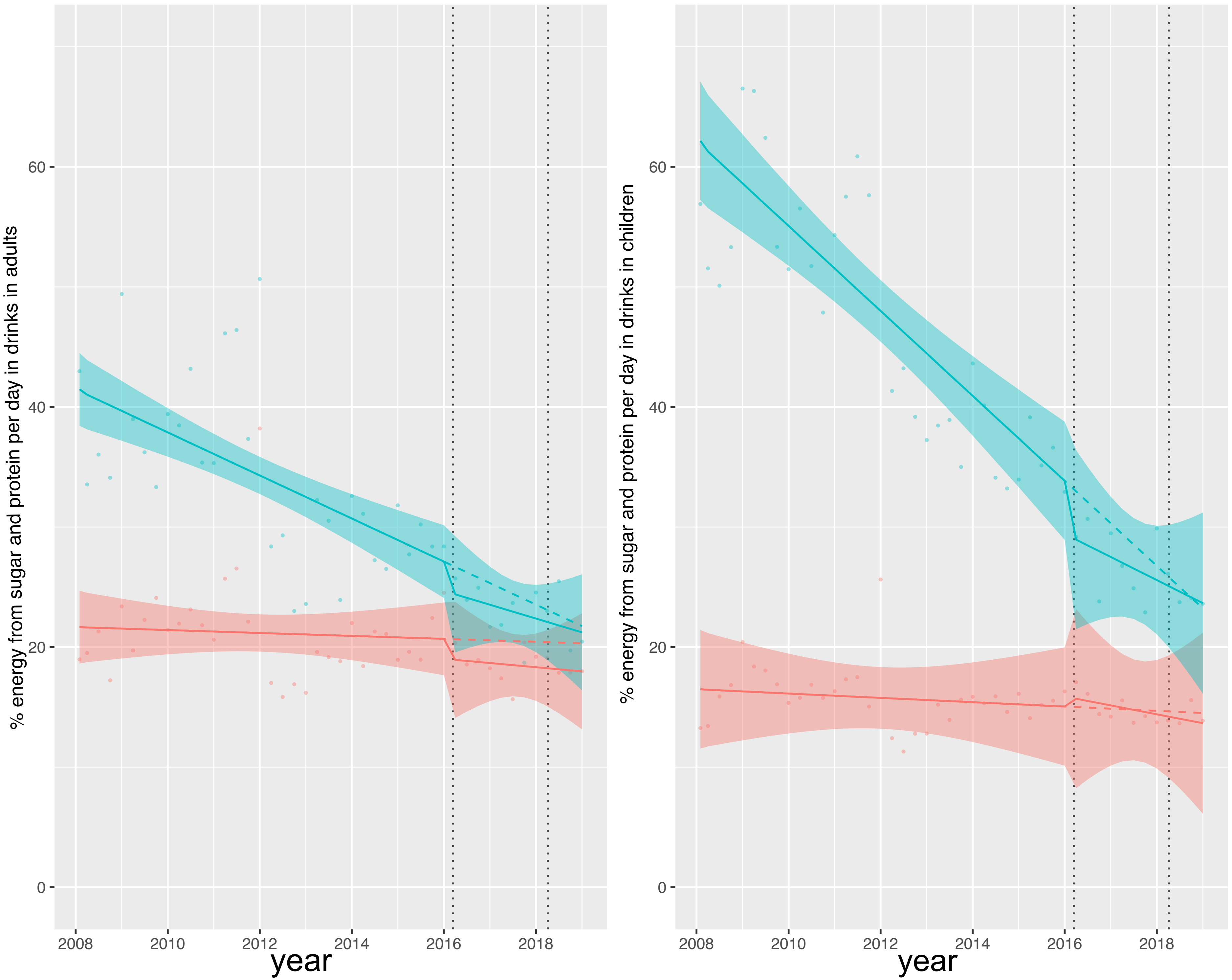

### Supplementary fig 4

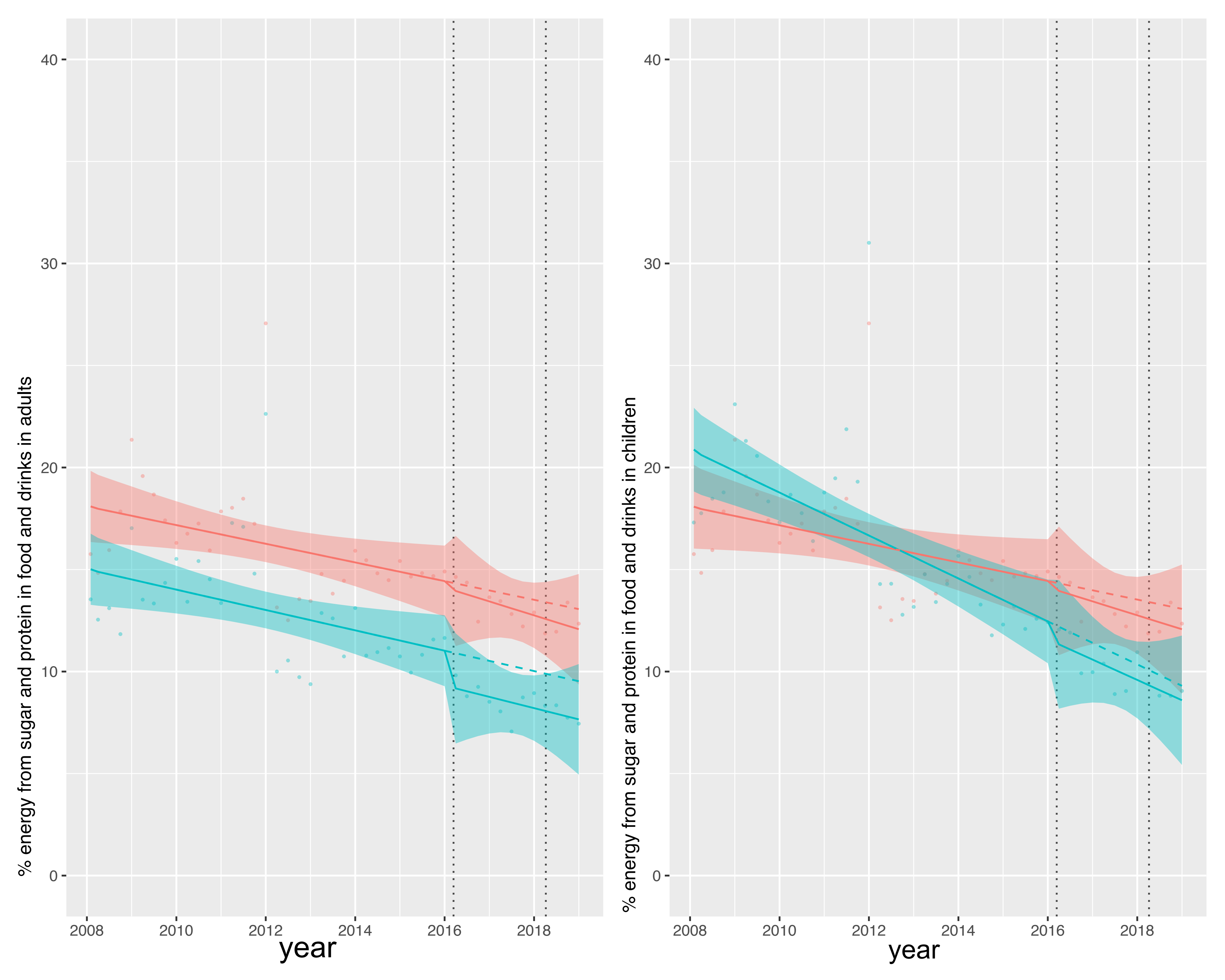
